# Modelling the synergy between single-occupancy and PPE in controlling COVID-19 outbreaks on hospital wards

**DOI:** 10.1101/2024.12.23.24319413

**Authors:** Cameron Zachreson, Robyn Schofield, Caroline Marshall, Marion Kainer, Kirsty Buising, Jason Monty, Sheena Sullivan, Kanta Subbarao, Nicholas Geard

## Abstract

**Background:** Outbreaks of respiratory pathogens on hospital wards present a major challenge for control of hospital-acquired infections. When illness presentation is mild or infection is asymptomatic, isolation of recognised cases may be insufficient to prevent outbreaks, as unrecognised cases may be common. In such scenarios, structural controls such as the design of wards with single-occupancy patient rooms, or routine precautions such as the use of N95 respirators by healthcare staff can play an important role in preventing and mitigating outbreaks.

**Methods:** This study applies an agent-based extension of the Wells-Riley model of airborne pathogen exposure to simulate COVID-19 outbreaks on hospital wards. We simulated the impact of single-vs. double-occupancy patient rooms on secondary attack rates and the sizes of outbreaks resulting from introduction of unrecognised cases. We further simulated the impact of N95 respirator use by nurses during patient care activities, assuming an efficacy of 90% for protection and source control.

**Results:** In our simulations, the size of outbreaks recorded at day 14 was markedly lower in wards with only single-occupancy rooms, compared to double-occupancy rooms (with means of 15.2 and 25.1 infections, respectively). We found that nurses working on wards were more likely to acquire infection than patients. Higher patient room occupancy was associated with increased outbreak size, with a larger relative impact on patients than staff. N95 respirators were effective at mitigating outbreaks, with higher impacts in wards with single-occupancy patient rooms.

**Conclusions:** Single-occupancy rooms can greatly decrease the risk of hospital acquired airborne infection for patients. We show that single-occupancy hospital rooms can also reduce the number of healthcare workers infected during an outbreak of an airborne respiratory virus, but not to the same relative extent as patients. Due to the structural constraints limiting transmission between patients in different rooms, outbreaks were driven by transmission events involving nurses, which were effectively mitigated through the use of N95 respirators. Taken together, our results suggest that single-occupancy rooms are effective at reducing outbreak sizes. However, they are insufficient by themselves to prevent large outbreaks without mitigation efforts focused on limiting the potential for transmission involving healthcare workers, such as the use of N95 respirators.

## I. INTRODUCTION

Controlling the transmission of respiratory pathogens within hospital environments is an ongoing global challenge. Densely occupied healthcare settings create the conditions for large outbreaks of airborne pathogens among groups of people with risk factors for severe infection outcomes. Though transmission is preventable through the isolation or cohorting of infected people, respiratory pathogens often become contagious before the onset of symptoms, emphasizing the importance of infection control precautions implemented routinely and consistently, such as masking for source control and protection.

Outbreaks of SARS-CoV-2 on hospital wards have demonstrated highly heterogeneous outcomes, which appear to be sensitive to crowding, the use of personal protective equipment (PPE) by healthcare workers, and room occupancy [1–5]. Even when patients are effectively protected through routine PPE use, contact tracing, and isolation of recognised cases, outbreaks can still propagate among healthcare workers due to asymptomatic transmission from unrecognised cases [6–8].

The structural design of hospital wards can influence transmission risk, and several studies have attempted to understand the potential for wards with primarily single-occupancy patient rooms to mitigate nosocomial acquisition of infections, with a focus on bacterial pathogens [9, 10]. While evidence generated by such studies has been mixed, observational studies of SARS-CoV-2 outbreaks demonstrated that single-occupancy wards exhibited dramatically reduced risk of large outbreaks [4]. The example of COVID-19 illustrates the critical challenge of controlling pathogen spread for diseases with highly heterogeneous illness presentation. When transmission occurs while infected individuals are asymptomatic, the capacity to control transmission through case isolation is greatly reduced. In such scenarios, infection control through the structural design of wards may be critically important to outbreak mitigation.

During the height of the COVID-19 pandemic, face coverings such as N95 respirators were used to prevent transmission, and their efficacy was subject to broad consensus, based on strong observational evidence that N95 respirators mitigate transmission [11–13]. After extensive vaccination campaigns targeting healthcare workers and during the transition to endemicity, the use of face coverings has declined.

This work investigates the individual and combined effects of structural controls and N95 respirators as standard practice, applied regardless of clinical presentation of infectious disease. Because of the non-linear nature of disease transmission the presence or absence of other controls may alter the marginal impact of additional measures [14, 15]. Here we examine coupled mitigation effects of single-occupancy rooms and N95 respirator use, implemented as standard practice (i.e., in effect regardless of infection status). In this case, our results demonstrate that these two control measures enhance one-another, with single-occupancy rooms increasing the benefit of N95 respirator use.

## II. METHODS

We applied an agent-based extension of the Wells-Riley model of airborne pathogen exposure to simulate COVID-19 outbreaks on hospital wards. We simulated the impact of single-versus double-occupancy patient rooms on secondary attack rates and outbreak size. We further simulated the impact of N95 respirator use by nurses during patient care activities, assuming an efficacy of 90% for both protection and source control.

### A. Airborne pathogen transmission

Structurally, the simulated ward consists of patient rooms (double-or single-occupancy) and a break room where nurses go for meals and short breaks during their shifts (Figure 1a). Pathogen transmission is computed as a stochastic process governed by Poisson statistics, with infection rate proportional to the concentration of viral quanta as in the Wells-Riley model of airborne pathogen exposure (Figure 1a, inset). Briefly, infected individuals shed virus into the space they occupy. Simultaneously, virus is removed through ventilation (specified in air changes per hour, ACH). If an individual who is susceptible to infection enters the contaminated zone, they are exposed to the viral concentration *c* during the period Δ*t* over which they occupy it. For such an exposure event, the probability of becoming infected is computed as:

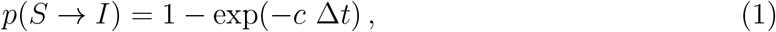

in which *S* → *I* denotes the transition from the susceptible to infected state. In our implementation we compute *c* for all individuals every eight hours by aggregating the exposure levels and time intervals corresponding their activity schedules, which are updated every five minutes (Figure 1b). The transient dynamics of viral concentration in each room due to ventilation is computed through a fine-grained numerical integration at a time scale of one minute. This multi-timescale approach allows our model to account for fluctuations in viral concentration while also achieving a higher level of computational efficiency when evaluating infection probabilities. See the Supplemental Material for more details about model implementation.

**FIG 1.**
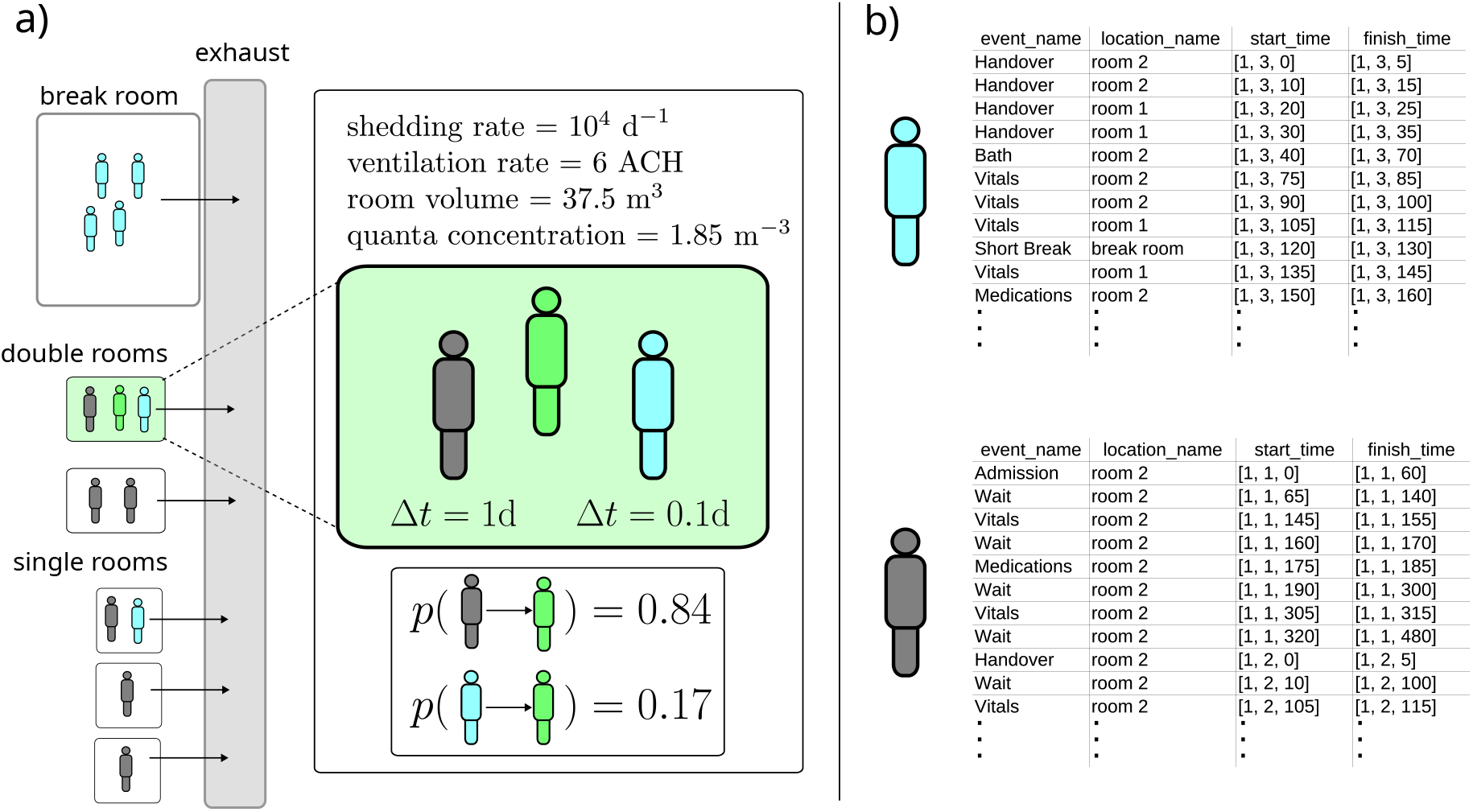
Schematic of model elements used to simulate airborne pathogen transmission in hospitals. Subfigure (a) shows a simple example of the structural model alongside a demonstration of exposure calculations for infection probability. In (a), green figures represent infected individuals, green-shaded rooms correspond to areas contaminated with virus, teal figures represent nurses, and gray figures represent patients. Subfigure (b) provides examples of activity trajectories for nurses (top) and patients (bottom), each activity is characterised by a name, a location, a start time, and a finish time.

### B. Ward structure

Throughout, we assume that double-occupancy rooms are 1.5 times the volume of single-occupancy rooms, which is a low-end estimate based on 2018 standards for modern Australian hospitals [16]. Transmission dynamics are initiated by introducing multiple infected patients (two or four, depending on the scenario). Because of our focus on standard precautions for infection control (as opposed to outbreak response), we restrict the duration of our simulations to 14 days. We further assume for simplicity that the patient population is fixed throughout (no discharges or new admissions). We assume mechanical ventilation inlets are located in rooms and exhaust vents are located in the corridors connecting them. This arrangement means we assume negligible mixing of air between patient rooms and between patient rooms and the break room used by nurses.

### C. Activity model

The activity model simulates the movement of nurses between different locations on the ward. It also accounts for patient length-of-stay, and the roster schedule determining which nurses are present for each 8-hr shift.

We assume that patients are attended to at least three times per 8-hr shift, once for medications (for a duration of 10 min) and twice for recording of vital signs (10 min per visit). Patients are also bathed once per day (30 min). We assume admission of patients takes one hour (but recall that in the scenarios investigated here the patient population is fixed, so admission and discharge do not play a major role). At the start of each 8-hour shift, handover of patients between nurses working subsequent shifts occurs (five minutes per patient).

Nurses each attend to four beds, which are organised into sections. For double-occupancy wards, sections span two rooms with two patients each. In addition to attending to patients, nurses take two short breaks per shift (10 min each) and one long break (60 min). Over a 14-day roster period, each nurse works a maximum of eight shifts, with no more than six of these performed on consecutive days. Consecutive shifts fewer than 16 hours apart are not allowed (i.e., multiple shifts within a 24-hour period are not allowed). Full details of the activity model for hospital wards are provided in the Supplemental Material.

### D. Numerical experiments

Our study consists of three types of numerical experiments, each corresponding to a specific question we aim to address:

1. How does single-occupancy affect transmission?
2. How does single-occupancy affect outbreak size?
3. How does single-occupancy affect the impact of N95 respirator use by nurses?

The corresponding experiments are described in Table I which provides the model variables that change in each scenario and the simulation output that is measured from each. We first examine the secondary attack rate from infected patients in a small ward with only four patients and six nurses. Then, we examine outbreaks in larger wards (40 patients, 53 nurses) with varied room occupancy and use of N95 respirators by nurses. For additional information on the impact of N95 respirator use by nurses in the context of single-or double-occupancy patient rooms, we evaluate each scenario for increasing values of pathogen infectiousness (controlled by the parameter ⟨*β*⟩, see Supplemental Material). Observing the size of outbreaks for increasing pathogen infectiousness allows us to qualitatively evaluate the impact of room occupancy and respirator use, independently and in combination, on the effective reproductive ratio.

**TABLE I.** Summary of numerical experiments performed in this study. SAR: mean secondary attack rate over an ensemble of simulations (average number of infections produced by the index cases); ⟨*I*_tot_⟩: mean total infections after 14 days over an ensemble of simulations (in our results, infection totals are further stratified between nurses and patients).

| question | beds | nurses | beds<br>per<br>room | mask<br>policy | no.<br>index<br>cases | output | notes |
| --- | --- | --- | --- | --- | --- | --- | --- |
| 1 | 4 | 6 | 1 | none | 2 | SAR | transmission from<br>secondary cases is<br>disabled<br>($\langle\beta\rangle = 10^4$) |
| 1 | 4 | 6 | 2 | none | 2 | SAR | transmission from<br>secondary cases is<br>disabled<br>($\langle\beta\rangle = 10^4$) |
| 2 | 40 | 53 | 1 | none | 4 | $\langle I_{\text{tot}} \rangle$ | final size<br>distribution for a<br>single<br>infectiousness<br>value ($\langle\beta\rangle = 10^4$) |
| 2 | 40 | 53 | 2 | none | 4 | $\langle I_{\text{tot}} \rangle$ | final size<br>distribution for a<br>single<br>infectiousness<br>value ($\langle\beta\rangle = 10^4$) |
| 3 | 40 | 53 | 1 | none | 4 | $\langle I_{\text{tot}} \rangle$ | systematically<br>varying<br>infectiousness<br>($500 \geq \langle\beta\rangle \leq 10^5$) |
| 3 | 40 | 53 | 2 | none | 4 | $\langle I_{\text{tot}} \rangle$ | systematically<br>varying<br>infectiousness<br>($500 \geq \langle\beta\rangle \leq 10^5$) |
| 3 | 40 | 53 | 1 | N95<br>(nurses) | 4 | $\langle I_{\text{tot}} \rangle$ | systematically<br>varying<br>infectiousness<br>($500 \geq \langle\beta\rangle \leq 10^5$) |
| 3 | 40 | 53 | 2 | N95<br>(nurses) | 4 | $\langle I_{\text{tot}} \rangle$ | systematically<br>varying<br>infectiousness<br>($500 \geq \langle\beta\rangle \leq 10^5$) |

## III. RESULTS

### A. Secondary attack rates for wards with single or double occupancy

We begin by characterising the model’s behaviour over a single generation of transmission. That is, we initialise the transmission dynamics by introducing index cases, and count the number of secondary infections they produce. We disable viral shedding from secondary cases to ensure we only count infections produced by the first generation of transmission. By counting infections in patients and nurses separately, we can distinguish the role of behavioural heterogeneity in the transmission process which will aid our interpretation of results from simulations of multi-generation outbreaks.

In our model, an infected patient in a single-occupancy room has no capacity to transmit the pathogen to any other patients. However, they may still infect healthcare workers. For double-occupancy rooms, an infected patient may transmit to the other patient sharing the same room, or to a healthcare worker caring for either of the room’s occupants.

To illustrate the effects of single-occupancy rooms on the potential for a patient to transmit infection, we simulate a small-scale ward with only four beds and six nurses. Due to the constraints imposed by our behavioural model, this provides a scenario in which only one nurse is present on the ward at any given time. We investigate three configurations of patient rooms and the initial placement of index cases (illustrated in Figure 2a). In the first of these, two infected patients are placed into single-occupancy rooms, which completely prevents transmission between patients. In the second, two infected patients are introduced and placed into the same double-occupancy room, which also prevents transmission between patients but changes the exposure level for nurses. In the third scenario, the two infected patients are each placed into different double-occupancy rooms, allowing for up to two secondary cases in patients. Distributions of secondary case numbers from 5000 independent simulations are shown for nurses in Figure 2b and for patients in Figure 2c.

**FIG 2.**
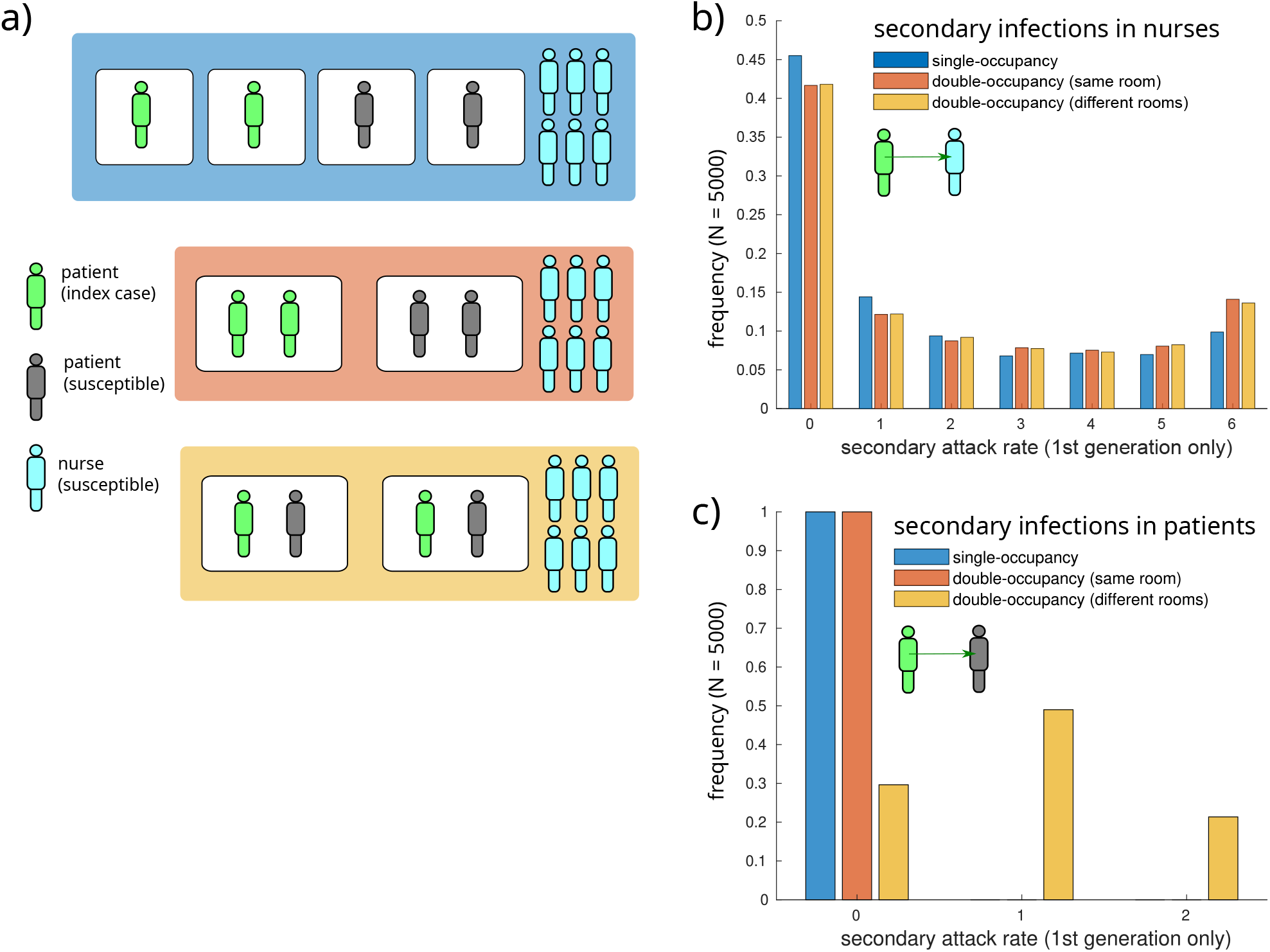
Single-occupancy rooms reduce secondary infections among both patients and nurses. Three scenarios are shown, each with a different configuration of index cases arranged into single-or double-occupancy rooms. Schematics of these scenarios are shown in (a). In the schematics, index cases are colored green, susceptible patients are gray, and susceptible nurses are teal. Boxes around patients represent rooms, while the larger colored boxes indicate correspondence between each scenario and the bar color in histograms (b) and (c) which illustrate secondary infection rates in nurses and patients, respectively, after a single generation of transmission. For the histograms in (b) and (c), 5000 simulations were conducted, with the maximum viral shedding rate of index cases sampled from a Gamma distribution, and all other simulation parameters held fixed (See methods).

Due to the relatively rapid changeover of healthcare workers on a hospital ward (three ward nurses per section per day, organised into shifts), the initially infected patients may transmit to many (up to six) healthcare workers over the course of their infectious periods (Figure 2a). While this qualitative difference is not sensitive to room occupancy, the probability of a patient infecting all six healthcare workers increases substantially for double-occupancy rooms (Figure 2a). This occurs because, while the viral concentration in a double room containing only one infected patient is lower than it would be in a single-occupancy room, the nurse servicing that room spends twice as much time there because they attend to both patients. Summary statistics for the distributions shown in Figure 2b and 2c are shown in Table II.

**TABLE II.**
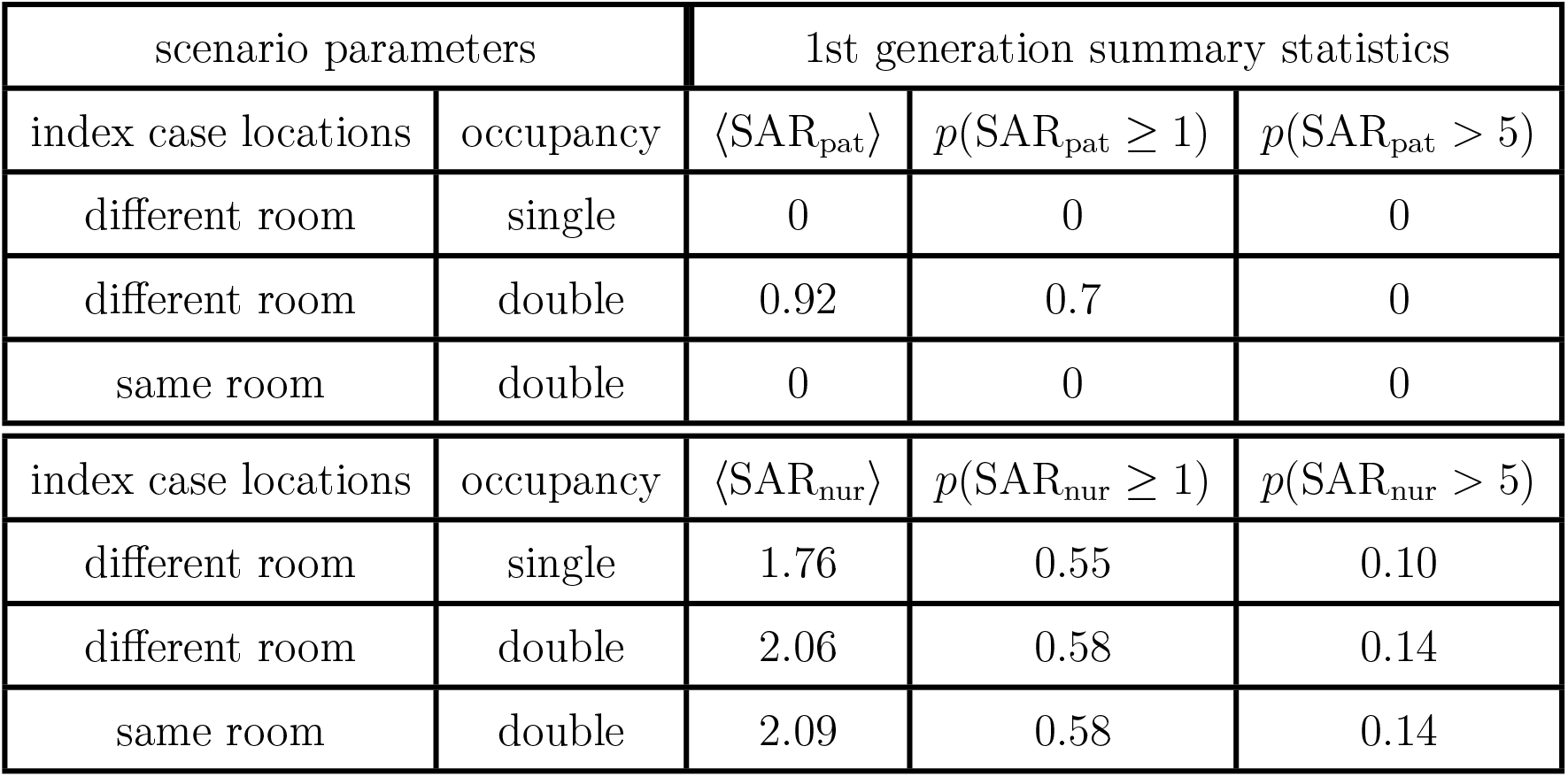
Summary statistics for secondary infection rates in patients and nurses for a four-bed ward with two initially infected patients.

| scenario parameters |  | 1st generation summary statistics |  |  |
| --- | --- | --- | --- | --- |
| index case locations | occupancy | $\langle \text{SAR}_{\text{pat}} \rangle$ | $p(\text{SAR}_{\text{pat}} \geq 1)$ | $p(\text{SAR}_{\text{pat}} > 5)$ |
| different room | single | 0 | 0 | 0 |
| different room | double | 0.92 | 0.7 | 0 |
| same room | double | 0 | 0 | 0 |
| index case locations | occupancy | $\langle \text{SAR}_{\text{nur}} \rangle$ | $p(\text{SAR}_{\text{nur}} \geq 1)$ | $p(\text{SAR}_{\text{nur}} > 5)$ |
| different room | single | 1.76 | 0.55 | 0.10 |
| different room | double | 2.06 | 0.58 | 0.14 |
| same room | double | 2.09 | 0.58 | 0.14 |

Our modelling approach produces an equivalence between exposure time and the concentration of airborne quanta, which is apparent when comparing the two different double-occupancy scenarios. Even though they have different positioning of index cases, the secondary attack rate in nurses is the same for both (Figure 2b, yellow and orange bars).

### B. Infection rates for unmitigated outbreaks

To understand the role that single-occupancy rooms may play in controlling the size of outbreaks, we performed simulations with a realistic ward size, over a 14-day time window, without N95 respirators. Here, the ward contained 40 patient beds, arranged into single-or double-occupancy rooms, with a total healthcare workforce of 53 nurses (10 nurses staff the ward at any given time). For simplicity, we assumed that all beds were occupied and the patient population was fixed over the simulation, not allowing discharge or admission of new patients. Schematics of these scenario configurations are shown in Figure 3a, with distributions of total infection numbers shown as histograms in Figures 3b and 3c.

**FIG 3.**
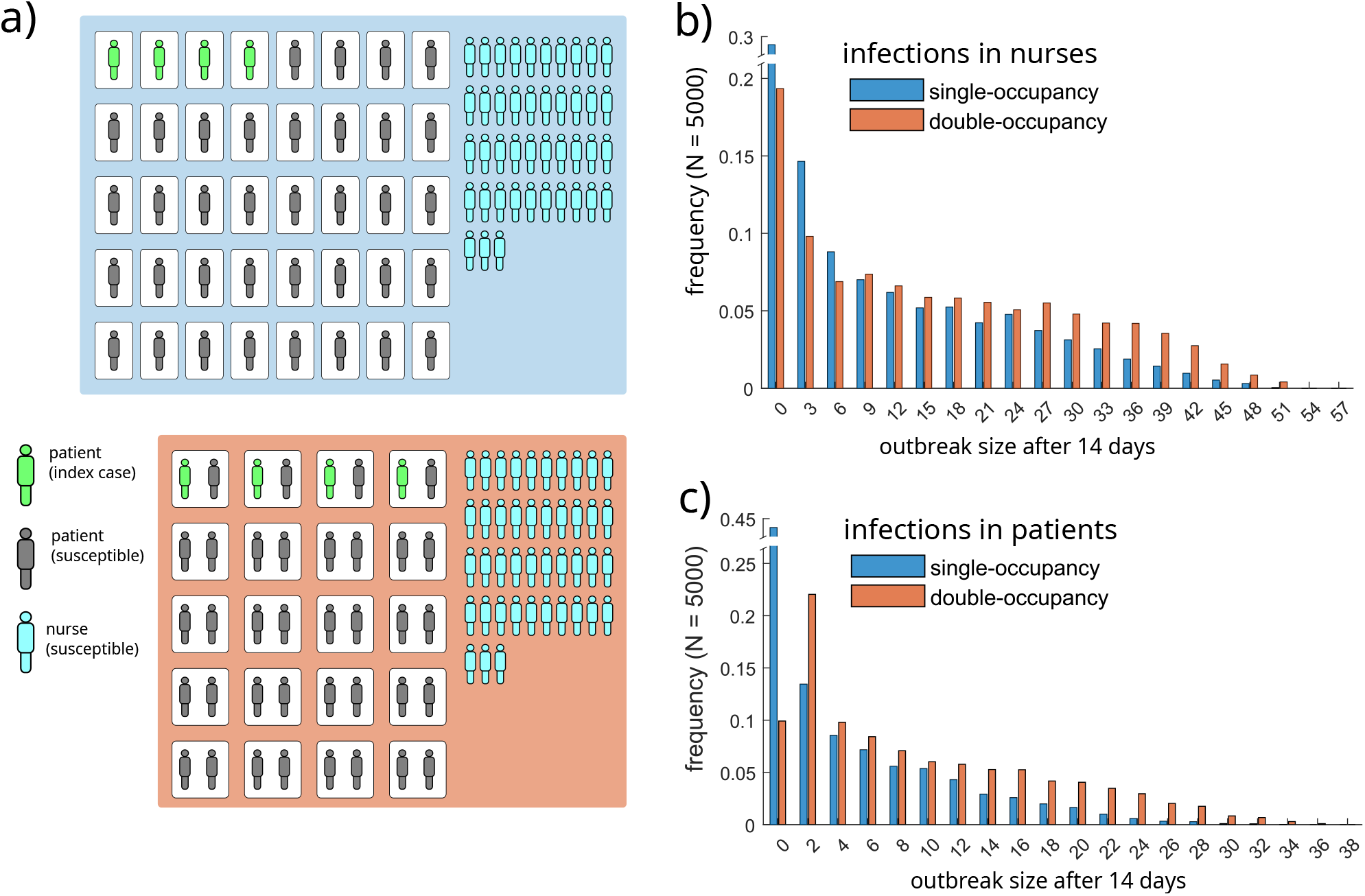
Single occupancy rooms reduce the size of outbreaks. Two scenarios are shown, each initialised with four infected patients. Schematics of these scenarios are shown in (a), with index cases colored in green, susceptible patients in gray, and susceptible nurses in teal. White boxes around patients represent rooms, while the larger colored boxes indicate correspondence between each scenario and the bar color in histograms (b) and (c) which total infection numbers in nurses and patients, respectively, after 14 days. For the histograms in (b) and (c), 5000 simulations were conducted, with the maximum viral shedding rate of index cases sampled from a Gamma distribution, index cases randomly assigned to rooms, and all other simulation parameters held fixed (See methods). Note the split y-axis scales in (b) and (c).

For both patients and nurses, outbreaks in wards with single-occupancy rooms produced substantially fewer infections. With double-occupancy rooms, the mean number of total infections after 14 days was 25.1, which decreased to 15.2 for single-occupancy wards. The absolute change was similar for nurses and patients, with mean total infections decreasing from 9.0 to 4.3 (patients), and from 16.1 to 10.9 (nurses). Additional summary statistics are shown in Table III.

**TABLE III.** Summary statistics for total infections after 14 days in patients and nurses for a 40-bed ward with four initially infected patients.

| scenario parameters | outbreak summary statistics |  |  |
| --- | --- | --- | --- |
| occupancy | $\langle I_{\text{patients}} \rangle$ | $p(I_{\text{patients}} \geq 1)$ | $p(I_{\text{patients}} > 10)$ |
| single | 4.3 | 0.56 | 0.16 |
| double | 9.0 | 0.9 | 0.37 |
| occupancy | $\langle I_{\text{nurses}} \rangle$ | $p(I_{\text{nurses}} \geq 1)$ | $p(I_{\text{nurses}} > 10)$ |
| single | 10.9 | 0.80 | 0.4 |
| double | 16.2 | 0.86 | 0.57 |

We note that while the impact of single-occupancy on secondary infection rates in nurses was relatively small (Figure 2b), the impact on final infection numbers was substantial (Figure 3b). This result highlights the indirect impact of mitigating transmission between patients on the overall outbreak dynamics. On the other hand, our results also illustrate the ongoing risk to patients posed by infections among nurses, even when transmission between patients is mitigated with single-occupancy rooms.

### C. Effect of N95 respirators on infection rates

Here, we investigate the potential for use of N95 respirators by healthcare workers to further mitigate outbreaks. We assume N95 respirators are used by healthcare workers when they are performing tasks in patient rooms. During breaks, no masks are used. N95 Respirators are assumed to reduce inhaled virus by 90% (if worn by a susceptible healthcare worker) and to reduce exhaled virus by 90% (if worn by an infected healthcare worker). Our simulation approach follows that which was described in the previous section except that here, we examine systematically increasing values of pathogen infectiousness. This allows us to observe (to a coarse approximation) the effects of occupancy and mask wearing on the reproductive ratio of the pathogen in the simulated context.

The use of N95 respirators substantially mitigates outbreaks, regardless of room occupancy (Figure 4). When they are not used (Figure 4a), outbreak sizes begin to sharply increase as pathogen infectiousness exceeds ⟨*β*⟩ values between 2500 and 5000. When N95 respirators are used, outbreaks remain small until ⟨*β*⟩ exceeds 10^4^, indicating a substantial decrease in the basic reproductive ratio. Single-occupancy rooms increased the impact of masking on total infections after 14 days (Tables IV).

**TABLE IV.** Effect of N95 respirator use and single-occupancy rooms on the mean final size of simulated outbreaks.

| | $\langle I_{\text{tot}} \rangle$ | | | | | |
| --- | --- | --- | --- | --- | --- | --- |
|  | single-occupancy |  |  | double-occupancy |  |  |
| $\langle \beta \rangle$ | N95 resp. | no N95 resp. | $\Delta$ (%) | N95 resp | no N95 resp. | $\Delta$ (%) |
| 500 | 0.037 | 0.460 | -92.0 | 0.783 | 1.396 | -43.9 |
| 1000 | 0.099 | 0.940 | -89.5 | 1.046 | 2.371 | -55.9 |
| 2500 | 0.255 | 2.767 | -90.8 | 1.696 | 6.164 | -72.5 |
| 5000 | 0.612 | 6.987 | -91.2 | 2.479 | 11.995 | -79.3 |
| 10000 | 1.724 | 15.981 | -89.2 | 4.121 | 24.047 | -82.9 |
| 25000 | 6.088 | 38.961 | -84.4 | 12.665 | 53.381 | -76.3 |
| 50000 | 16.373 | 61.066 | -73.2 | 26.332 | 71.507 | -63.2 |
| 100000 | 30.982 | 73.164 | -57.7 | 46.641 | 81.031 | -42.4 |

**FIG 4.**
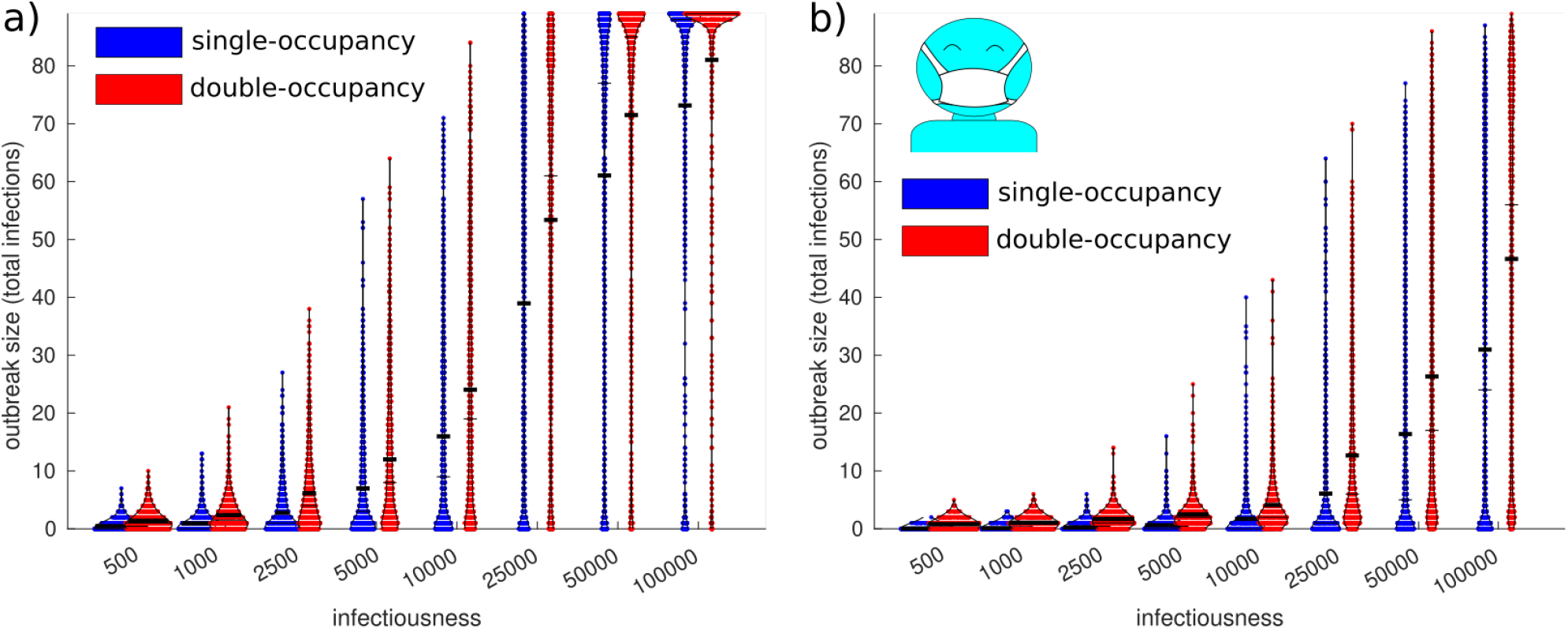
The use of N95 respirators by nurses reduces outbreak sizes, particularly in combination with single-occupancy rooms, across a range of values for pathogen infectiousness. Swarm plots show the distribution of final size values for each scenario of occupancy and respirator use. Subfigure (a) corresponds to the baseline condition with no use of respirators while (b) corresponds to the use of respirators by healthcare workers while treating patients in their rooms (we assume no use of respirators during breaks). Red points correspond to wards with only double-occupancy patient rooms while blue dots correspond to wards with only single-occupancy rooms. Thick horizontal lines correspond to sample means, while thin horizontal lines correspond to medians. The x-axis values correspond to the parameter ⟨*β*⟩ which controls the infectiousness of the pathogen (see Methods).

For infections in patients, the impact of masking monotonically decreased with pathogen infectiousness for single-occupancy rooms because direct transmission between patients is not possible (Table V). For double-occupancy rooms, the impact of nurses wearing N95 respirators on patient infections increases from a relative change of -2.7% to -83.9% for 500 ≤ ⟨*β*⟩ ≤ 2.5 *×* 10^4^ and then decreases for higher values of pathogen infectiousness. This is because, for low levels of contagiousness, the pathogen cannot transmit effectively between patients and nurses (regardless of mask use) so a small number of transmission events between patients is responsible for most of the infections. As contagiousness increases, the potential for transmission between nurses and patients begins to play a substantial role in outbreak dynamics, and N95 respirators play a greater role in mitigation. As contagiousness increases further, the impact of respirator use decreases because (even with 90% efficacy) they become less effective at curbing outbreaks.

**TABLE V.**
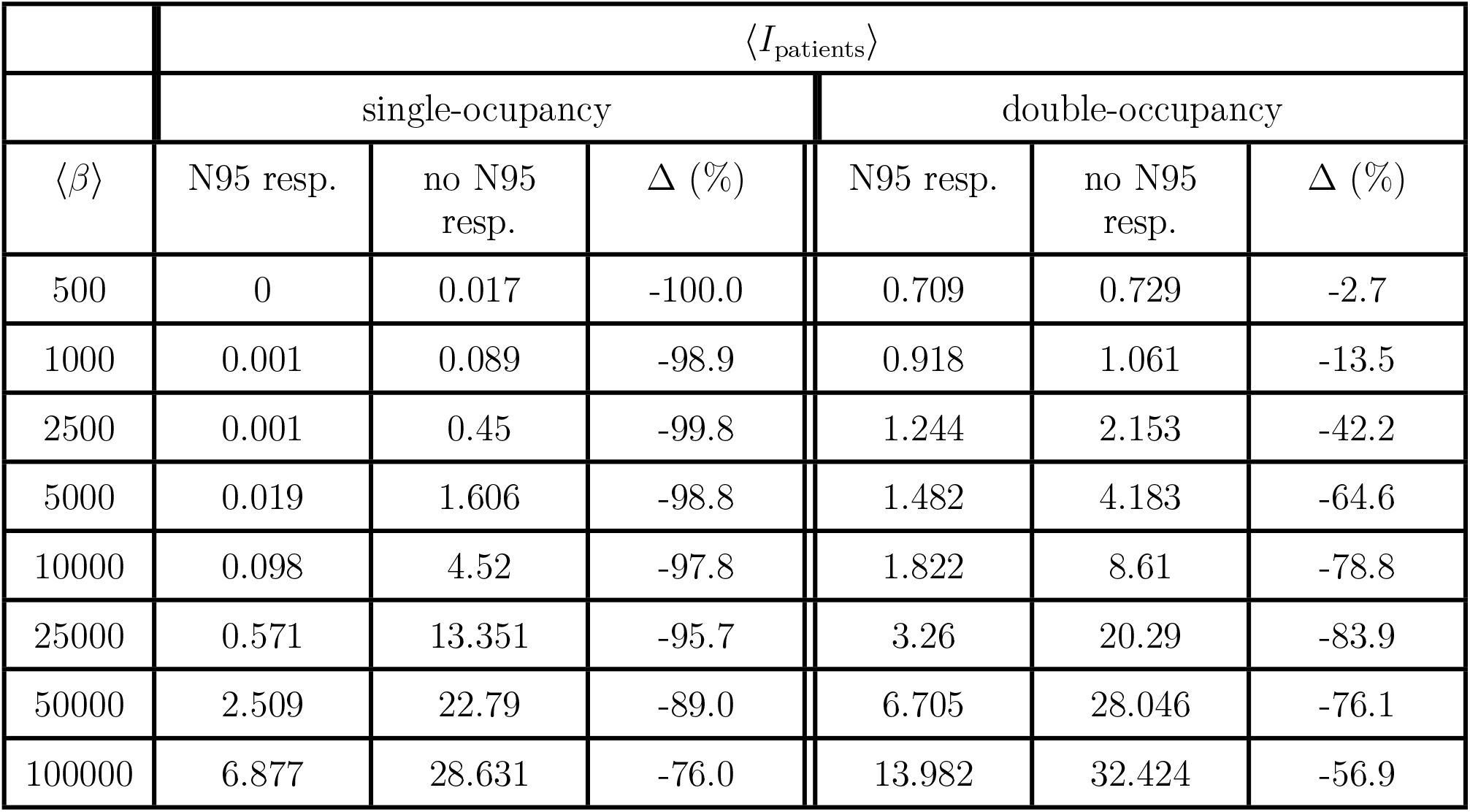
Effect of N95 respirator use and single-occupancy rooms on the mean number of infected patients after 14 days.

| | $\langle I_{\text{patients}} \rangle$ | | | | | |
| --- | --- | --- | --- | --- | --- | --- |
|  | single-occupancy |  |  | double-occupancy |  |  |
| $\langle \beta \rangle$ | N95 resp. | no N95 resp. | $\Delta$ (%) | N95 resp. | no N95 resp. | $\Delta$ (%) |
| 500 | 0 | 0.017 | -100.0 | 0.709 | 0.729 | -2.7 |
| 1000 | 0.001 | 0.089 | -98.9 | 0.918 | 1.061 | -13.5 |
| 2500 | 0.001 | 0.45 | -99.8 | 1.244 | 2.153 | -42.2 |
| 5000 | 0.019 | 1.606 | -98.8 | 1.482 | 4.183 | -64.6 |
| 10000 | 0.098 | 4.52 | -97.8 | 1.822 | 8.61 | -78.8 |
| 25000 | 0.571 | 13.351 | -95.7 | 3.26 | 20.29 | -83.9 |
| 50000 | 2.509 | 22.79 | -89.0 | 6.705 | 28.046 | -76.1 |
| 100000 | 6.877 | 28.631 | -76.0 | 13.982 | 32.424 | -56.9 |

With respect to infections in nurses, the use of N95 respirators plays a substantial role regardless of room occupancy because it directly prevents them from being infected by patients. However, the impact is again higher for single-occupancy than it is for double-occupancy, with more noticeable effects of room occupancy for higher pathogen contagiousness (Table VI).

**TABLE VI.** Effect of N95 respirator use and single-occupancy rooms on the mean number of infected nurses after 14 days.

| | $\langle I_{\text{nurses}} \rangle$ | | | | | |
| --- | --- | --- | --- | --- | --- | --- |
|  | single-occupancy |  |  | double-occupancy |  |  |
| $\langle \beta \rangle$ | N95 resp. | no N95 resp. | $\Delta$ (%) | N95 resp. | no N95 resp. | $\Delta$ (%) |
| 500 | 0.037 | 0.443 | -91.6 | 0.074 | 0.667 | -88.9 |
| 1000 | 0.098 | 0.851 | -88.5 | 0.128 | 1.31 | -90.2 |
| 2500 | 0.254 | 2.317 | -89.0 | 0.452 | 4.011 | -88.7 |
| 5000 | 0.593 | 5.381 | -89.0 | 0.997 | 7.812 | -87.2 |
| 10000 | 1.626 | 11.461 | -85.8 | 2.299 | 15.437 | -85.1 |
| 25000 | 5.517 | 25.61 | -78.5 | 9.405 | 33.091 | -71.6 |
| 50000 | 13.864 | 38.276 | -63.8 | 19.627 | 43.461 | -54.8 |
| 100000 | 24.105 | 44.533 | -45.9 | 32.659 | 48.607 | -32.8 |

## IV. DISCUSSION

The confluence of risk factors in healthcare settings presents unique challenges for infection control. In hospitals, vulnerable patients with high prevalence of acute health conditions and chronic comorbidities have high rates of close contact with co-located patients and healthcare staff. Additionally, during the 24-hr roster cycle of a hospital ward, the high rate of exchange of healthcare workers produces a continuous level of indirect contact with the outside world. As such, the compounding risks of i) exposure to infectious pathogens and ii) adverse clinical outcomes of infection are both amplified, relative to normal social conditions. Healthcare environments in well-resourced settings often have the capacity to isolate patients using negative-pressure spaces to prevent transmission of airborne pathogens. However, successful isolation of infected patients who do not present with infection-associated illness upon admission requires continuous disease surveillance, which is costly both in terms of time and resources.

Some pathogens have consistent presentation of symptoms alongside infectiousness, which makes outbreaks easier to control [17]. For example, early containment of the SARS pandemic in early 2003 was feasible in part due to the tendency for severe illness presentation alongside infectiousness. Similarly, transmission of Ebola virus is typically mediated by direct contact with symptomatic cases [18]. However, for pathogens such as Influenza and SARS-CoV-2, disease presentation is highly heterogeneous and large fractions of infectious cases present mild illness or are completely asymptomatic at the time of transmission [19–21].

Since the emergence of SARS-CoV-2 as a pandemic disease, outbreaks in healthcare facilities such as hospitals and residential care facilities have been common. Due to the high rate of transmission due to unrecognised cases, transmission-based precautions for respiratory infection control have become an important aspect of healthcare provision. Such precautions may prevent transmission of SARS-CoV-2 between patients and healthcare workers, and are broadly considered helpful for preventing outbreaks [11].

The use of N95 respirators for source control has been demonstrated to reduce viral shedding into the air, and there is substantial observational evidence for the effectiveness of face coverings at preventing transmission [12, 13]. However, due to the difficulty of designing and conducting high quality randomised controlled studies in healthcare environments, evidence for direct estimates of the overall efficacy of N95 respirators has been assessed as insufficient [13, 22]. Structural design features of hospital wards can also have dramatic impacts on the ability to control airborne pathogen outbreaks. In the context of COVID-19, observational studies have identified room occupancy as a key risk factor for transmission between patients [4, 23].

Largely due to their potential to mitigate pathogen transmission, there has been a shift towards the design of hospitals with primarily single-occupancy rooms in some parts of the world [24]. While single-occupancy hospitals may carry higher initial costs, they are broadly considered advantageous in terms of patient well-being [25], infection control [4, 9, 23, 26, 27], and operating costs [25]. Furthermore, surveys of healthcare workers have shown favorable attitudes among staff in single-occupancy hospitals, with patient observation challenges arising as a the most significant drawback [26, 28, 29]. By demonstrating how physical separation between patients can increase the benefit of PPE use by healthcare staff, our results support the design of hospital wards with single-occupancy rooms.

## V. CONCLUSIONS

In this work, we used a purpose-built agent based model to simulate COVID-19 outbreaks on hospital wards. Through simulations, we investigated the combined and marginal impacts of single-occupancy rooms and the use of N95 respirators by healthcare workers as standard precautions. We found that the benefits of these layered infection control strategies were synergistic: single-occupancy rooms reduced outbreak sizes and also increased the impact of N95 respirator use. Intuitively, this synergy results from the distinct impacts of the two infection control features: single-occupancy rooms reduce transmission between patients, while N95 respirators reduce transmission between patients and healthcare workers. Neither intervention targets transmission between healthcare workers when they are not caring for patients and therefore not wearing N95 respirators (i.e., during breaks). Transmission between healthcare workers can be a major issue, as evidenced by large outbreaks among staff, even when outbreak control effectively prevents infections in patients [6–8]. As such, further investigations should focus on methods of preventing transmission between healthcare workers, such as increased air filtration and ventilation in shared spaces such as break rooms and nurses stations. Based on our present results, we expect such measures would offer additional synergistic effects, substantially reducing the size of outbreaks in hospital wards.

## Data Availability

All data produced are available online at https://github.com/cjzachreson/ward_model_public

## VI. ACKNOWLEDGEMENTS

This research was supported by the Medical Research Future Fund (MRFF) via a COVID-19 Treatment Access and Public Health Activities grant (Grant ID: MRF2017355). RS and JM are recipients of funding from ARC Industrial Transformation Training Centre (Grant ID: IC220100012).

## Supplemental Material

### S1. OVERVIEW

Our simulations of disease transmission caused by exposure to an airborne virus are based on the traditional Wells-Riley model[30, 31]. In the Wells-Riley model, exposure to an infectious pathogen is assumed to follow homogeneous Poisson statistics, in which the rate of infection is proportional to the concentration of virus to which an individual is exposed:

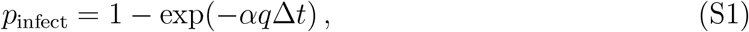

where *q* represents the concentration of infectious virus (quanta m^−3^), Δ*t* is the duration of time the susceptible individual is exposed, and the constant *α* absorbs contextual factors that may influence exposure such as the use of masks, breathing rate, or host-specific susceptibility. It is assumed in using Equation *S*1 that the concentration *q* is constant in time over the period *T*. In complex environments such as hospital wards, pathogen concentrations cannot be assumed constant for two reasons:

1. Ventilation rates are high, and involve a system of mechanical ventilation elements that create pressure differentials and move air between the different parts of the space.
2. Infected and susceptible individuals may be mobile, moving around between different parts of the ward as they shed and inhale airborne virus.

If either of the above conditions were not present, the simple exposure model expressed in Equation S1 would be sufficient to simulate disease transmission dynamics because a steady-state value for *q* could be calculated. However, because there is heterogeneity both in terms of the spatial configuration of ventilation, and in terms of the movement of individuals between spaces, we do not generally expect *q* to reach a steady state. To account for these processes, we take an agent-based simulation approach in which we specify *q* as a quantity that varies in space and time, and use the dynamic configuration of infected individuals to compute it for defined subregions of the space (i.e., different rooms). Then, we compute an individual-specific value of *p*_infect_ for each susceptible person by aggregating their exposure levels over their trajectory through the space. We then evaluate the infection status of susceptible individuals at a timescale commensurate with the infection process (more details below).

For the implementation used in this work, subregion zones correspond to patient rooms and the break room in which nurses eat and take short breaks. Our model has three time scales:

1. Δ*t*^(1)^ a coarse-grained time scale used to compute exposure, infection, disease progression and recovery,
2. Δ*t*^(2)^ a medium-grained time scale used to update the positions and activities of agents,
3. Δ*t*^(3)^ a fine-grained time scale used to compute the concentration of virus in each space of the hospital ward.

We update the locations (zones) occupied by individuals every five minutes (the behavioural timescale Δ*t*^(2)^), use a fine-grained forward-Euler approach to compute the dispersion of virus via airflow between zones (the airflow timescale Δ*t*^(3)^), and aggregate exposure every 8hr (exposure timescale Δ*t*^(1)^) to compute and evaluate *p*_infect_ for each individual *i*. Because we implement our model in discrete time, we ensure that Δ*t*^(1)^ = *n*_1_Δ*t*^(2)^ and Δ*t*^(2)^ = *n*_2_Δ*t*^(1)^ where *n*_1_ and *n*_2_ are positive integers.

To express this approach precisely, we first partition our model into two main layers: the **Ward Model** (WM) and the **Transmission Dynamics Model** (TDM). The WM describes the spatiotemporal dynamics of patient and healthcare worker behaviour and its output is a comprehensive list of all events which occur on a ward. The TDM includes within-host disease progression, viral shedding from infected agents, exposure of susceptible agents to virus, and a networked airflow model which describes how airborne viral contamination disperses around the ward.

In our modelling approach, the WM generates a set of agent trajectories which is then used as input to the TDM to simulate outbreaks. This sequential layered approach produces the limitation that agent behaviours which alter the timing or positioning of activities are not sensitive to outbreak dynamics (i.e., preventive physical distancing cannot be explicitly simulated). However, it offers a number of advantages including reduced runtime and RAM usage, and the capacity to efficiently create experimental hierarchies (for example, modifying components of the TDM while controlling all aspects of the WM). Furthermore, splitting the WM and the TDM into separate software components will allow the use of the same TDM for completely different applications beyond hospital wards (any behavioural model that produces a set of trajectories with a compatible format can be used as input). Below, we provide full details of the WM and TDM layers:

### S2. WARD MODEL

To simulate the behaviour of nurses and patients on a hospital ward, the model uses the following conceptual approach: nurses attend the ward on a rotating roster (details below) and are assigned to groups of beds (sections) that can be occupied by patients receiving care. Nurses are required to take breaks, and to service the basic health needs of patients. When a patient is admitted to the ward they are assigned to an available bed and the essential care tasks associated with that patient are added to a queue. When the nurse assigned to the patient’s bed is available (not currently performing a task), they initiate the next task in the queue. The dynamics of the ward play out as nurses finish tasks and start new ones. Finished tasks are tabulated and registered into an output which records a trajectory of events for each agent, these agent trajectories are the principal output of the WM which is fed into the TDM. More details are provided below:

The WM defines the following high-level structures:

- Ward: defines the spatial and temporal structure of the ward environment
- Entities: objects included in the simulation such as patients, nurses, beds, and rooms. Entities have an integer-valued ID number.
- Event: a discrete event describing a place, time interval, and subset of nurses and patients involved
- Agent Trajectory: the sequence of events involving a specific patient or nurse description notes

**TABLE S1.** The elements of a simulated ward.

| object name | description | notes |
| --- | --- | --- |
| Nurses | map {nurse ID → Nurse} | all nurse entities indexed by integer ID |
| Admitted Patients | map {patient ID → Patient} | patients currently present on ward, indexed by integer ID |
| Roster | map {shift ID → [nurse ID]} | links each roster shift (via shift ID) to an array of nurse IDs |
| Shifts | map {shift ID → Shift} | links each shift ID to a Shift object |
| Bed sections | map {Section ID → Section} | links each Section ID to a Section object |
| Beds | map {Bed ID → Bed} | links each Bed ID to a Bed object |

#### A. Ward structure

A Ward object is the high-level container that organises the various WM elements. It has internal structures corresponding to the sets of nurse and patient Entities, the hospital beds and their respective assignments to staffing sections and rooms, and a schedule to determine which nurses are present in the ward on a given shift. The key elements of a Ward object are listed in Table S1.

The Ward’s characteristics include maps that index ID numbers to the various entities present on the ward. These include patients, nurses, beds, bed sections (assigned to nurses), the roster assigning nurses to shifts, and the shifts describing the assignment of nurses to bed sections. A comprehensive list of Entities and their main properties is provided in Table S2.

**TABLE S2.** Entities included within a simulated ward.

| Entity | property | description |
| --- | --- | --- |
| Nurse |  |  |
|  | section ID | (Int) ID of the group of beds assigned to the nurse |
|  | break schedule | (Array of Events) timing and duration of breaks |
|  | worked last shift | (Bool) True if nurse worked previous shift |
|  | worked shift before last | (Bool) True if nurse worked shift before last |
|  | n consecutive shifts | (Int) number of consecutive days with any shift |
|  | n shifts this roster | (Int) total number of shifts during a roster cycle |
|  | is eligible | (Bool) indicates if nurse is eligible to be assigned a shift |
|  | is busy | (Bool) indicates if a nurse is available to start a new task |
| Patient |  |  |
|  | bed ID | (Int) index of a patient's assigned bed |
|  | length of stay | (Int) number of shifts between admission and discharge |
| | service schedule | map $\{(\text{day ID}, \text{shift ID}) \rightarrow [\text{Events}]\}$<br>links a day and a shift to a set of healthcare events |
| Bed |  |  |
|  | room ID | (Int) ID of the room containing the bed |
|  | section ID | (Int) ID of group of beds (assignable to nurse) |
|  | is occupied | (Bool) indicates if bed is allocated to a patient |
|  | is assigned | (Bool) indicates if bed is assigned to a nurse |
|  | patient ID | (Int) ID of patient allocated to bed |
|  | nurse IDs | (Int-valued set) IDs of any nurses assigned to bed |
| Shift |  |  |
|  | time of day | (String) "morning", "daytime" or "nighttime" |
|  | day index | (Int) ID of day containing the shift |
| | Section assignments | map $\{\text{section ID} \rightarrow [\text{Nurse IDs}]\}$<br>and reverse map $\{\text{Nurse ID} \rightarrow [\text{Section ID}]\}$<br>links nurse IDs to section IDs and vice versa |

#### B. Generating a roster

The WM configures a Ward object and then uses it to generate a trajectory of all the events that occur on the ward over a specified duration. The most complex aspect of configuring a Ward object is generation of a Roster that reflects a given set of workforce constraints and practices. As indicated in Table S1, a Roster object describes which nurses work on which shifts throughout the simulated time period. It is organised into segments describing roster cycles which reflects the organisational strategy used by hospitals in Australia. In a given roster cycle, there are several rules that constrain which nurses are eligible to be assigned a given shift:

1. the total number of shifts a nurse is already assigned in a given roster cycle,
2. the number of consecutive days a nurse has worked,
3. whether or not the nurse has been assigned in one of the previous two shifts.

Building a valid roster involves iterating through the days (14 per cycle) shifts (three per day) of each roster cycle and assigning eligible nurses to bed sections that are not yet assigned. Once all bed sections have been assigned for a given shift, the roster generation process moves to the next shift and repeats the assignment process after updating the eligibility status of each nurse. To prevent the iterative roster generation process from failing (i.e., running out of eligible nurses before all shifts in a roster cycle have been filled), assignment of nurses to unfilled bed sections preferences nurses who have a lower workload in the given roster cycle. This not only prevents the algorithm from failing, but also ensures that the workload is evenly distributed.

#### C. Generating agent activity trajectories

After building a roster, synthetic trajectories are generated based on patient care schedules, nurse break schedules, and nurse availability. Some care activities have scheduled start times within a shift, ensuring that sufficient time elapses between (e.g.) medication delivery and the recording of vital signs. Other activities (bathing, admission, discharge) are not given scheduled start times and are carried out whenever a nurse is available.

Within each 8-hr shift, activities on the ward are simulated in discrete time, in increments of five minutes. A queue of activities is first generated for the shift based on scheduled breaks and patient care requirements. Trajectories are built by iterating forward in time and initiating each activity when the conditions arise that it can be carried out (when it is scheduled, and/or when a nurse is available to perform the activity). Within each 5-min time step, a list is generated containing the IDs of all nurses who are available to undertake new tasks. If any of those nurses have a break scheduled in the current time step, those activities are added to a queue of next events. Then, events involving patients are added to the queue of next events. Events that are currently underway progress forward by 5-min and, if their set duration is reached, are added to a growing list of finished events. A comprehensive list of the activities which take place on the ward is provided in Table S3. One example each of a patient’s activity trajectory and a nurse’s activity trajectory over a single 8-hr shift are provided in Tables S4 and S5, respectively. Note the format used for activity times corresponds to [day index, shift index, minute of shift].

**TABLE S3.**
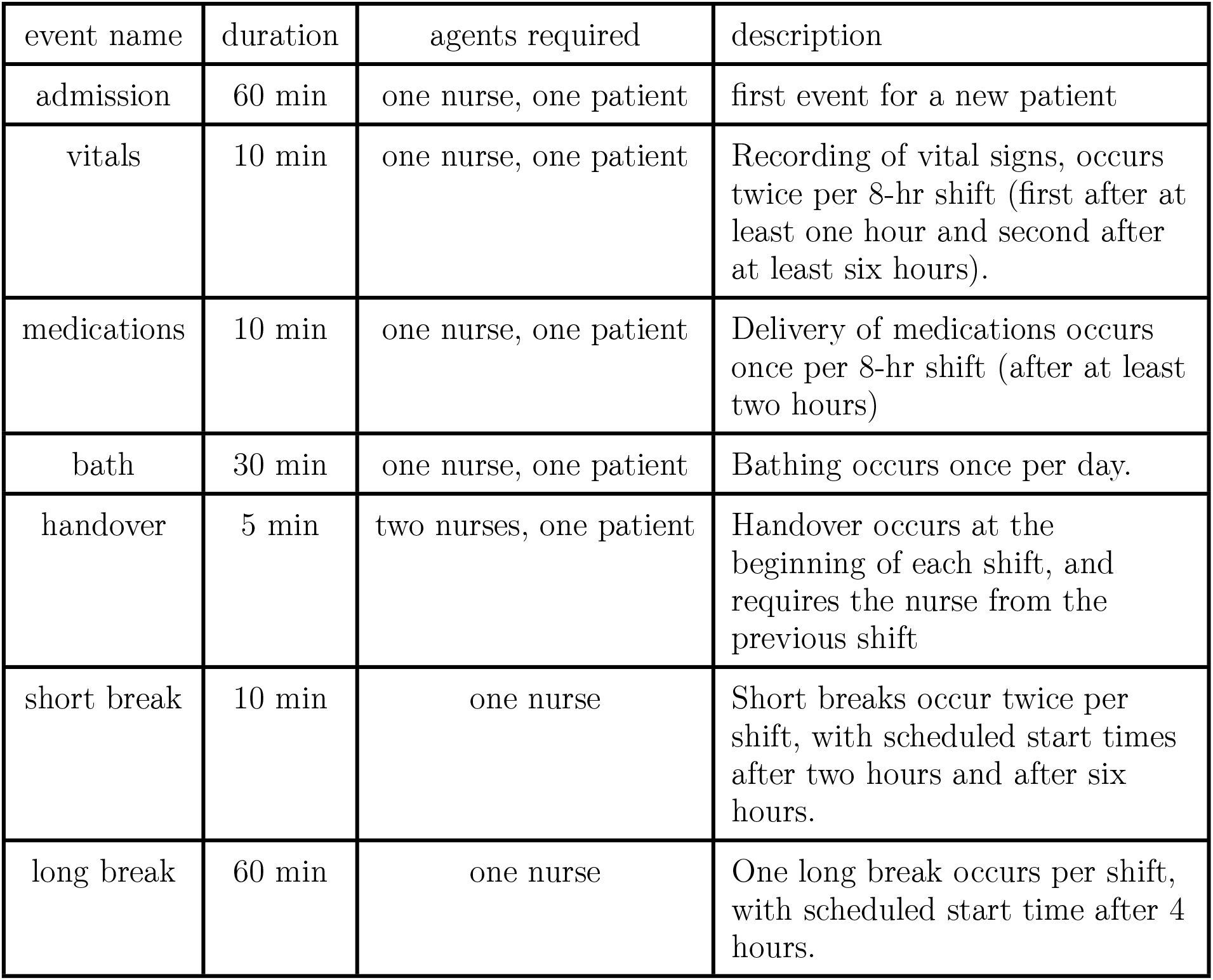
Activities that take place on a simulated ward.

**TABLE S4.**
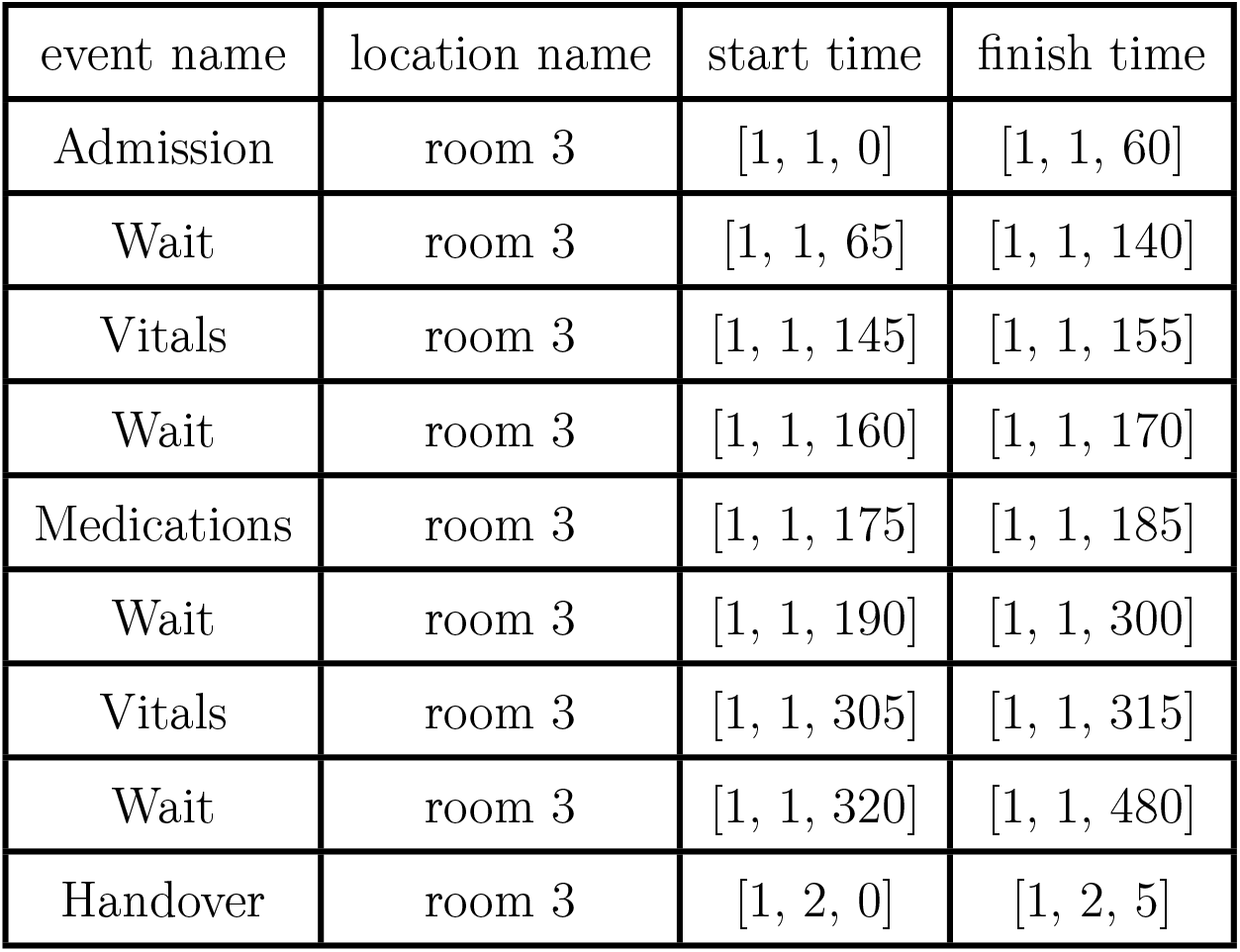
The activities of a single patient over one 8-hr shift.

**TABLE S5.**
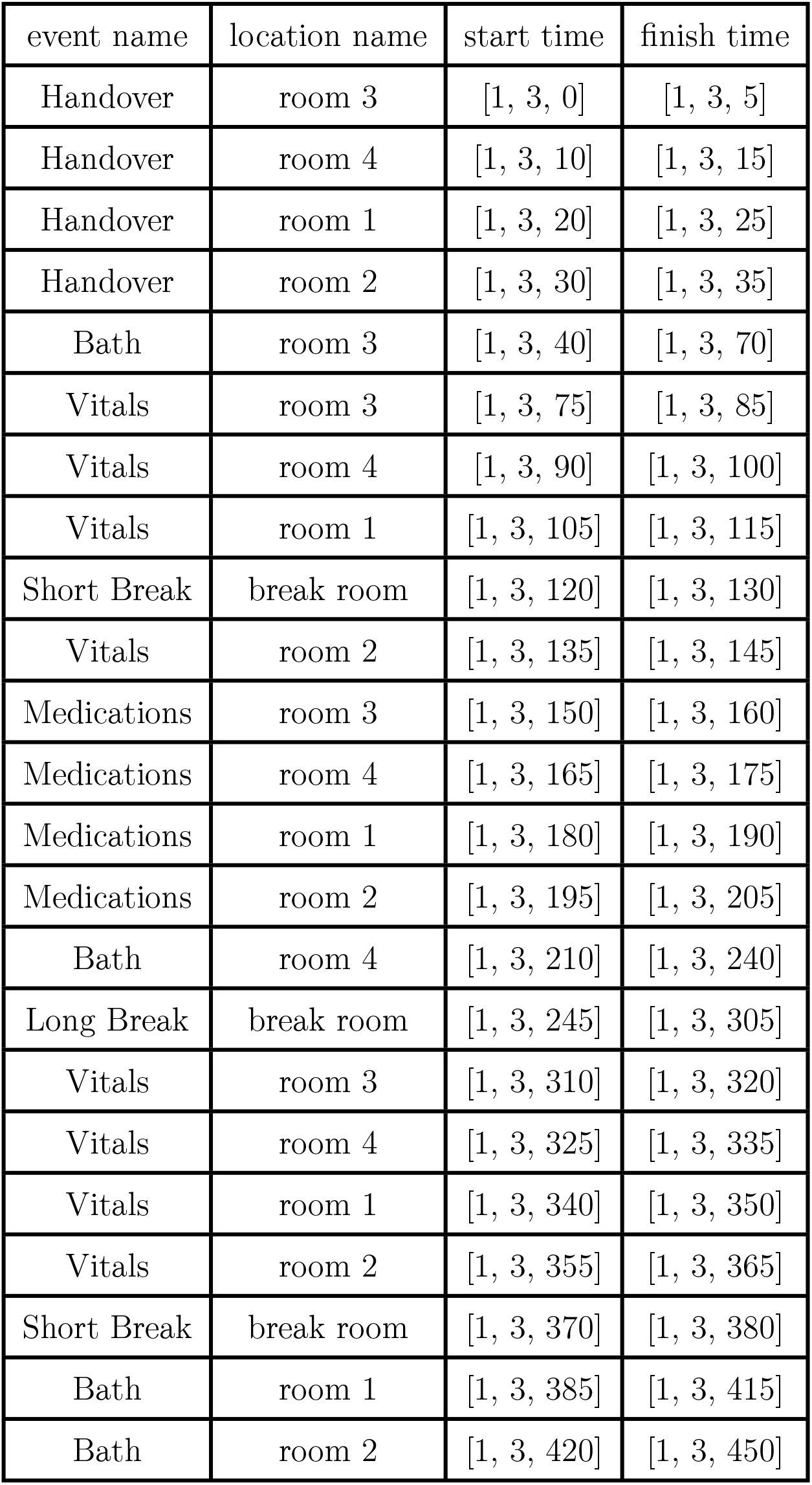
Activities undertaken by one nurse during an 8-hr shift.

| event name | location name | start time | finish time |
| --- | --- | --- | --- |
| Handover | room 3 | [1, 3, 0] | [1, 3, 5] |
| Handover | room 4 | [1, 3, 10] | [1, 3, 15] |
| Handover | room 1 | [1, 3, 20] | [1, 3, 25] |
| Handover | room 2 | [1, 3, 30] | [1, 3, 35] |
| Bath | room 3 | [1, 3, 40] | [1, 3, 70] |
| Vitals | room 3 | [1, 3, 75] | [1, 3, 85] |
| Vitals | room 4 | [1, 3, 90] | [1, 3, 100] |
| Vitals | room 1 | [1, 3, 105] | [1, 3, 115] |
| Short Break | break room | [1, 3, 120] | [1, 3, 130] |
| Vitals | room 2 | [1, 3, 135] | [1, 3, 145] |
| Medications | room 3 | [1, 3, 150] | [1, 3, 160] |
| Medications | room 4 | [1, 3, 165] | [1, 3, 175] |
| Medications | room 1 | [1, 3, 180] | [1, 3, 190] |
| Medications | room 2 | [1, 3, 195] | [1, 3, 205] |
| Bath | room 4 | [1, 3, 210] | [1, 3, 240] |
| Long Break | break room | [1, 3, 245] | [1, 3, 305] |
| Vitals | room 3 | [1, 3, 310] | [1, 3, 320] |
| Vitals | room 4 | [1, 3, 325] | [1, 3, 335] |
| Vitals | room 1 | [1, 3, 340] | [1, 3, 350] |
| Vitals | room 2 | [1, 3, 355] | [1, 3, 365] |
| Short Break | break room | [1, 3, 370] | [1, 3, 380] |
| Bath | room 1 | [1, 3, 385] | [1, 3, 415] |
| Bath | room 2 | [1, 3, 420] | [1, 3, 450] |

### S3. TRANSMISSION DYNAMICS MODEL

Abstractly, our extension of the Wells-Riley model can be described as a discrete-time stochastic Susceptible, Infected, Recovered (SIR) simulation of disease transmission. The probability of each susceptible agent becoming infected over the *k*-th time interval is computed as:

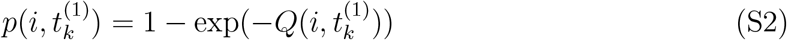

where *Q* has units of viral quanta days per cubic meter and represents the mean rate of infection for susceptible individual *i* over the time interval (*t*_*k*_ − Δ*t*^(1)^, *t*_*k*_]. The infection probability 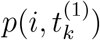 is updated on the ordered list of *n* discrete simulation time steps *t*^(1)^ of duration Δ*t*^(1)^:

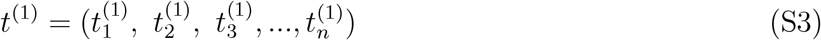

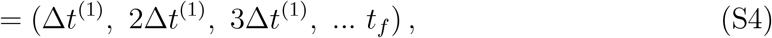

such that for each time step 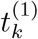, for each susceptible agent *i*, a Bernoulli trial is conducted with success probability 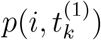 to determine if agent *i* becomes infected.

The rate 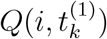 is computed from the finer-grained time scale Δ*t*^(2)^ as:

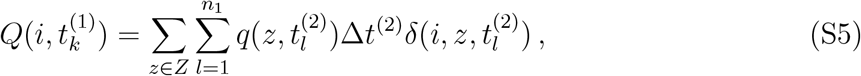

in which the set *Z* represents the set of zones with different viral concentrations, the indicator function 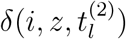 takes a value of 1 if agent *i* is located in zone *z* over the interval 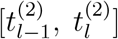 and takes a value of 0 otherwise, and the ordered list

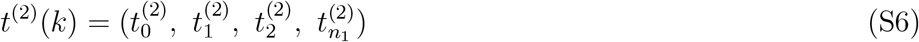

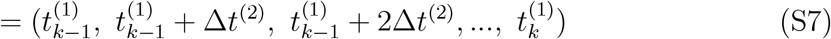

is a finer-grained set of discrete time steps with resolution Δ*t*^(2)^ = Δ*t*^(1)^*/n*_1_.

In words, Equation S5 for 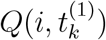 describes the aggregate exposure level of agent *i* to infectious virus particles over the time interval 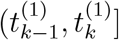 as the sum of the viral concentrations in each zone they visited over the period, weighted by the amount of time spent in each zone. In our model, the indicator function 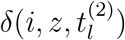 is determined by the trajectories produced by the WM (see previous section).

For each time step 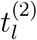, we compute the quanta concentration 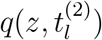 as a function of the locations and viral shedding rates of infected agents ***I***, the airflow between zones, and the concentration profile at the end of the previous time step 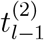:

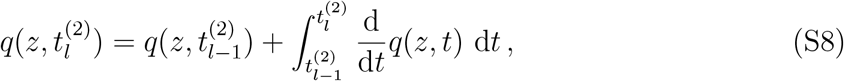

where the time-dependent concentration of virus in each zone *q*(*z, t*) depends on airflow patterns and the locations of infected people ***I*** in the zones *Z* via the following ODE :

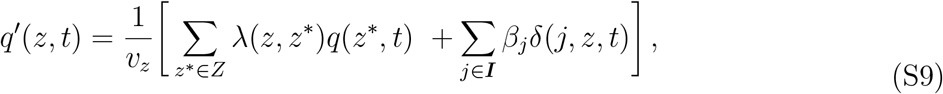

[Note that we use the notation *q*^*′*^(*z, t*) to represent the time derivative <u>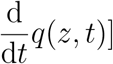</u>where *v*_*z*_ is the volume of zone *z, λ*(*z, z*^*^) is the air flow rate from zone *z* to zone *z*^*^ (volume per unit time), and *β*_*j*_ is the viral shedding rate of infected agent *j* (expressed in quanta per unit time). Again, the indicator function *δ*(*j, z, t*) represents the presence of agent *j* in zone *z* at time *t*.

We approximate 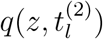 from Equation S9 as an initial value problem starting with the previous value 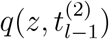 and iterating forward in time using forward-Euler step with the fine-grained time scale Δ*t*^(3)^:

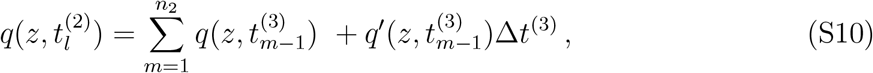

using the ordered list of fine-grained time steps

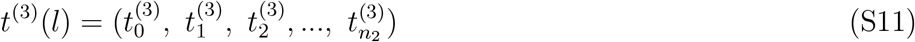

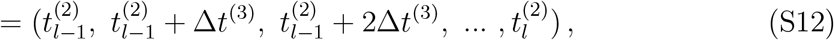

where that Δ*t*^(3)^ = Δ*t*^(2)^*/n*_2_.

#### A. Within-host model of disease progression

Note that the term *β*_*j*_ in Equation S9 is expressed as a constant. This is because we treat *β*_*j*_ as a constant on the time scale Δ*t*^(2)^. Between the time of infection *τ*_0_ and the time of recovery *τ*_rec_, the viral shedding rate for an infected agent *j* is updated at each step *τ* ∈ *t*^(1)^ :

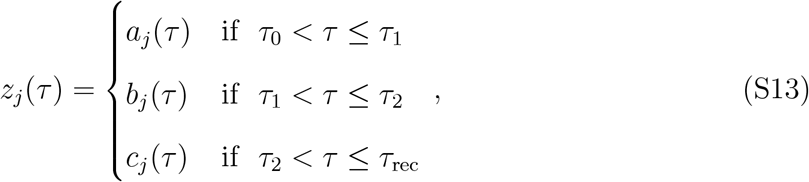

where *a*_*j*_(*τ*) describes the increase of infectiousness during the incubation period *t*_inc_, *b*_*j*_(*τ*) describes a brief plateau of infectiousness just before symptom onset (if the case is symptomatic), and *c*_*j*_(*τ*) describes the decline of infectiousness ending in recovery at time *τ*_recovery_. The time *τ*_0_ ∈ *t*^(1)^ corresponds to the time of infection, 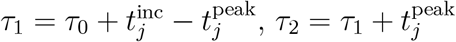, and 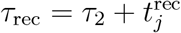. These segments of the piecewise function in Equation S13 are:

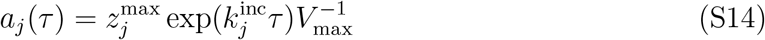

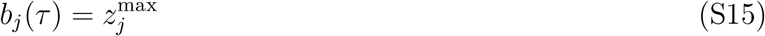

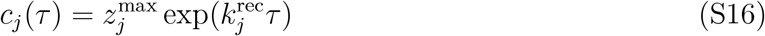

Finally, the function *z*_*j*_(*τ*) is re-scaled to calculate the time-dependent rate of infectious quanta production:

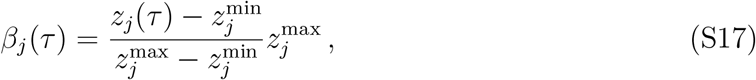

where 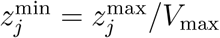. Equations S13 through S17, describe an initial exponential growth of infectious quanta, a plateau, followed by an exponential decline until recovery. For each infection, the scaling of these dynamics is determined by the individual-level parameters listed in Table S6 and the constant parameters listed in Table S7. More details about how this model was developed and calibrated for COVID-19 can be found in our previous work [14, 32]

**TABLE S6.** Individual-level parameters governing an infected person’s trajectory of infectiousness over time.

| symbol | value | description |
| --- | --- | --- |
| $z_j^{\max}$ | $z^{\max} \sim \text{Gamma}(\phi = 0.15, \langle\beta\rangle\phi^{-1})$ | peak (maximum) infectiousness |
| $t_j^{\text{inc}}$ | $t^{\text{inc}} \sim \text{Log-normal}(\mu = 1.62, \sigma = 0.418)$ | incubation period between infection and symptom onset (mean = 5.51 days) |
| $t_j^{\text{peak}}$ | $t_j^{\text{inc}} \Delta_{\text{peak}}$ | duration of infectiousness peak |
| $t_j^{\text{rec}}$ | $t^{\text{rec}} \sim \text{Uniform}(5, 10)$ | period between symptom onset and recovery |
| $k_j^{\text{inc}}$ | $\log(V_{\max})[t_j^{\text{inc}} - t_j^{\text{peak}}]$ | growth rate of infectiousness during incubation |
| $k_j^{\text{rec}}$ | $\log([V_{\max}]^{-1})[t_j^{\text{rec}}]^{-1}$ | decay rate of infectiousness during recovery (e.g., after onset of symptoms) |

### S4. NUMERICAL EXPERIMENTS

There are two types of numerical experiments that we used in our study. The first is designed to analyse a single generation of transmission, to assess the distribution of secondary cases produced by introducing infected patients to the ward environment. The objective of these single-generation experiments is to understand heterogeneity in exposure risk between patients and nurses on the ward. The second type of experiment assesses the size of an outbreak after a specified time horizon. Here, we use a time horizon of 14 days, to quantify the early dynamics of the outbreak. In both types of experiments, different ward configurations are investigated. In the second type of experiment (outbreaks) we also examine how the outbreak size changes with the constant ⟨*β*⟩, which modulates the mean infectiousness of the pathogen.

**TABLE S7.** Caption.

| symbol | value | description |
| --- | --- | --- |
| $\langle\beta\rangle$ | various ($10^4$ unless otherwise shown) | mean peak infectiousness |
| $V_{\max}$ | 7.0 | shape parameter controlling steepness of growth and decline of infectiousness |
| $\phi$ | 0.15 | shape parameter governing dispersion of infectiousness among agents |
| $\mu$ | 1.62 | log-mean of incubation period distribution |
| $\sigma$ | 0.418 | log-std of incubation period distribution |
| $\Delta_{\text{peak}}$ | 0.1 | fraction of incubation period over which infectiousness plateau lasts |

### A. Initialisation

#### 1. Activity trajectories

The agent trajectories are generated prior to simulating transmission dynamics, and are held fixed over the ensemble of simulation runs used for each experiment. Each ward configuration corresponds to a different set of trajectories, because the trajectories depend on the configuration of zones on the ward and the numbers of nurses and patients.

For the single-generation experiments, two different sets of trajectories are used, one for the single-occupancy ward layout and one for the double-occupancy layout. As discussed in the main text, these experiments use a small-scale model with only four patients (two or four rooms, depending on occupancy) and six nurses. For the outbreak simulations, a larger ward is used, with 40 patients and 53 nurses. Two different sets of trajectories are used, one for the single-occupancy ward layout, and one for the double-occupancy layout.

#### 2. Selecting index cases

In both types of experiment, initialisation specifies index cases - patients who will be infected at the beginning of the outbreak. These individuals are infected at the beginning of the simulation, with infections initialised at the first day after exposure (i.e., we assume they become infected while on the ward, and do not enter the ward after some period of infectiousness has already passed).

In single-generation experiments, two index cases are generated, and we investigate what happens if specific individuals appear as index cases (and whether or not they are sharing a room). As such, the index cases are selected specifically by their ID, in order to infect the patients who are in the locations desired for each scenario.

In outbreak experiments, four index cases are generated, selected at random from the patient population.

#### B. Simulation output

Each simulation produces two numbers, one corresponding to the number of nurses infected during the simulation, and the other corresponding to the number of patients infected. Each experiment performs a set number of instances, over which the ward configuration and agent trajectories are fixed (prior to initialisation), and the transmission dynamics are evaluated independently at random (using a different seed to initialise the pseudo-random number generator for each instance). Results are analysed as summary statistics produced from the ensemble associated with each experiment.

